# Prognostic Impact of Preoperative Atrial Fibrillation in Patients Undergoing Heart Surgery in Cardiogenic Shock

**DOI:** 10.1101/2023.06.15.23291469

**Authors:** Mariusz Kowalewski, Giuseppe M. Raffa, Michał Pasierski, Michał Pasierski, Michalina Kołodziejczak, Radosław Litwinowicz, Wojciech Wańha, Wojciech Wojakowski, Jan Rogowski, Marek Jasiński, Kazimierz Widenka, Tomasz Hirnle, Marek Deja, Krzysztof Bartus, Roberto Lorusso, Zdzisław Tobota, Bohdan Maruszewski, Piotr Suwalski, KROK Investigators

## Abstract

**Background:** Surgical intervention in the setting of cardiogenic shock (CS) is burdened with high mortality. Due to acute condition, detailed diagnoses and risk assessment is often precluded. Atrial fibrillation (AF) is a risk factor for perioperative complications and worse survival but little is known about AF patients operated in CS. Current analysis aimed to determine prognostic impact of preoperative AF in patients undergoing heart surgery in CS.

**Methods:** We analyzed data from the Polish National Registry of Cardiac Surgery (KROK) Procedures. Between 2012 and 2021, 4,852 patients presented with CS and were referred for cardiac surgery in 37 centers. Total of 624 (13%) patients had AF history. Cox proportional hazards models were used for computations. Propensity score (nearest neighbor) matching for the comparison of patients with and without AF was performed.

**Results:** Median follow-up was 4.6 years (max.10.0), mean age was 62 (±15) years and 68% patients were men. Thirty-day mortality was 36%. The origin of CS included acute myocardial infarction (36%), acute aortic dissection (22%) and valvular dysfunction (13%). In an unadjusted analysis, patients with underlying AF had almost 20% higher mortality risk (HR 1.19, 95% CIs 1.06-1.34; P=0.004). Propensity score matching returned 597 pairs with similar baseline characteristics; AF remained a significant prognostic factor for worse survival (HR 1.19, 95% CI 1.00-1.40; P=0.045).

**Conclusions:** Among patients with CS referred for cardiac surgery, history of AF was a significant risk factor for mortality. Role of concomitant AF ablation and/or left atrial appendage occlusion should be addressed in the future.

## Introduction

Cardiogenic shock (CS)-related condition at the time of cardiac surgery procedures is a common cause of mortality and its management remains a major challenge despite advances in therapeutic options including mechanical cardiovascular support (MCS)[1–3]. Some of the reversible causes of cardiogenic shock can be successfully managed surgically, provided they are diagnosed quickly before damage to the myocardium is permanent and recovery unlikely [4–6]. Regardless, cardiac surgery in patients with cardiogenic shock is often burdened with excessive risk [7–9], for the following reasons: 1) detailed diagnostic process might have been not performed due to extremely compromised patient hemodynamic condition; 2) surgery tends to focus on the main objective which is to reverse the CS with borderline coronary lesions or moderate valve insufficiencies seldom addressed; 3) risk of postoperative complications is much higher due to end-organ hypoperfusion and dysfunction at baseline; and finally; 4) postcardiotomy shock from low cardiac output syndrome (LCOS) is more likely to develop in these patients and postoperative MCS such as veno-arterial extracorporeal membrane oxygenation (V-A ECMO) or ventricle assist device (VAD) may be necessary alongside pharmacological support to stabilize the patients in this critical condition [10].

Atrial fibrillation (AF) is the most common arrhythmia worldwide and its prevalence is higher in patients with coronary artery-(CAD) or valve disease [11]. The effect of untreated AF on long-term prognosis, both in patients who need cardiac surgery and in patients who do not, is well known [12–14]. On the other hand, the available evidence on whether and how pre-existing AF is complicating cardiogenic shock is limited to acute myocardial infarction (AMI) induced CS (AMI-CS) [15–17] but poorly investigated in surgically treated CS patients. This is the first report to address the burden of AF in patients undergoing heart surgery for CS.

## Methods

Data were collected in a retrospective fashion from the KROK (Polish National Registry of Cardiac Surgery Procedures) registry (available at: www.krok.csioz.gov.pl). The registry is an ongoing, nationwide, multi-institutional registry of heart surgery procedures in Poland; the details on registry conception and design were described previously [18–20]. Study was approved by Institutional Board of CSK MSWiA and adheres to Helsinki Declaration as revised in 2013. Due to anonymization of registry data, patient consent was waived.

### Study population

The registry included all adult patients undergoing heart surgery between and 1^st^ Jan 2012 and 31^st^ Dec 2021 and presenting with cardiogenic shock due to all causes. Only patients undergoing heart surgery for isolated pericardial effusion were excluded. Cardiogenic shock in the KROK registry was defined as per SHOCK trial criteria [21] 1) systolic blood pressure (SBP) <90 mm Hg for >30 min or vasopressor support to maintain SBP >90 mm Hg or 2) evidence of end-organ damage (urine output [UO] <30 mL/h or cool extremities) or 3) hemodynamic criteria: cardiac index (CI) <2.2 and pulmonary capillary wedge pressure (PCWP) >15 mm Hg; until 2016; from then on, European Society of Cardiology Heart Failure guidelines [22] criteria were imposed and CS defined as 1) SBP <90 mm Hg with appropriate fluid resuscitation with clinical and laboratory evidence of end-organ damage or 2) clinical criteria: cold extremities, oliguria, altered mental status, narrow pulse pressure and 3) laboratory criteria: metabolic acidosis, elevated serum lactate, elevated serum creatinine.

Diagnosis of cardiogenic shock was left to discretion of treating physician. We divided the study cohort into patients with documented history of AF before the index surgery, and patients without documentation of AF. Post-operative AF was not recorded and therefore not considered. The study flow chart of the present analysis is shown in Figure 1.

**Figure 1.**
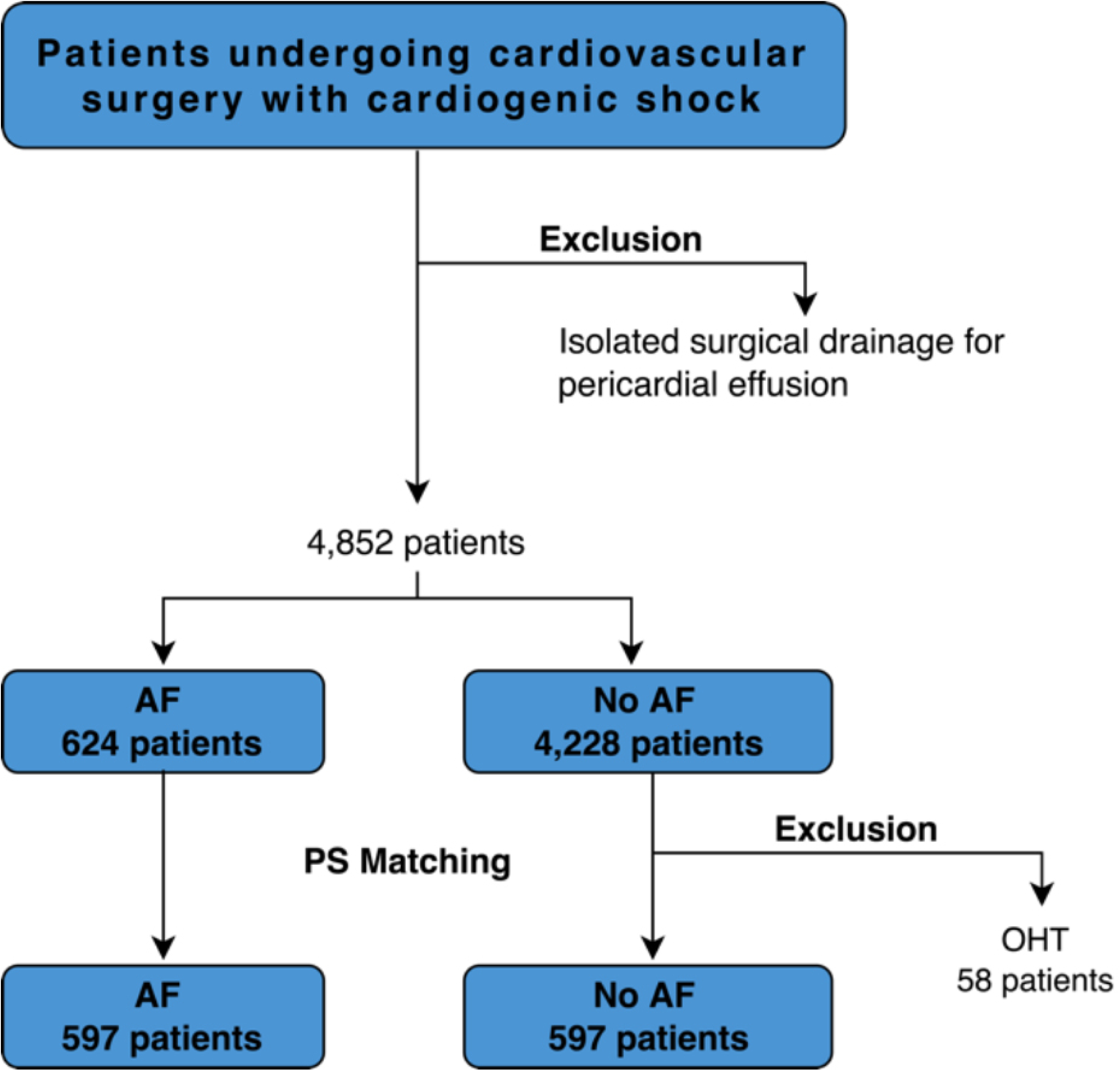
Study flow-chart. AF, atrial fibrillation; PS, propensity score; OHT, orthotopic heart transplantation.

### Clinical variables and endpoints

For patients undergoing heart surgery, we considered and reported 3 categories of variables: (1) baseline demographics: age, gender, EuroSCORE II [23] and its single components; (2) extent of coronary artery disease (CAD) and/or valvular and/or aortic disease and (3) surgical variables: urgency, operative technique (e.g. on-pump vs off-pump coronary artery bypass grafting [CABG] surgery). The primary endpoint was death from any cause reported at 30 days and longest available follow-up for the comparison of AF and non-AF patients. In-hospital outcomes and lengths of stays in the intensive care unit (ICU) and hospital (HLoS) are reported and compared as well. Baseline clinical-, procedural- and outcome data at follow-up were entered into prespecified electronic case report forms. Follow-up status with respect to all-cause mortality is validated by Polish National Health Fund and incorporated into the KROK registry.

### Statistical analysis

Registry records with >5% of missing data were not considered; in those with <5%, missing data were input by artificial neural networks [24]. Continuous variables were summarized as mean ± standard deviation if normally distributed; non-normal distributions were summarized as median and interquartile range (IQR) and compared with the Mann–Whitney U test or standard t test as appropriate. Categorical variables (number [%]) were compared with the Fisher’s exact test. Risk ratios (RRs) were used primarily for 30-day/in-hospital outcomes. Univariable and multivariable analyses to determine predictors of mortality were conducted. Similarly, we carried out univariable and multivariable analyses to identify the factors associated with the prevalence of AF. We built a non-parsimonious model including variables identified in multivariable analyses [25] for propensity score matching (PSM); a 1 to 1 nearest neighbor matching was performed with replacement (caliper 0.2); the overall long-term mortality was assessed with Kaplan-Meier curves fitted before (unadjusted model) and after propensity score matching. Cox regression was used to determine long-term hazard ratio (HR) for all-cause mortality as stratified by AF and non-AF patients. As a further sensitivity analysis to assess the survival in AF and non-AF subsets, we further stratified patients according to pre-defined subgroups. STATA MP v13.0 software (StataCorp, College Station, TX USA) and the packages “psmatch2”, “robust”, “optmatch”, “matchIt” and “CRTgeeDR” in R Core Team 2013 were used.

## Results

### Baseline demographics

Preoperative AF was documented in 624 of 4,852 (12.8%) patients, the mean age was 62 years and 68% patients were men. Baseline characteristics of unadjusted group of patients are further available as Supplementary Table 1. Presence of underlying atrial fibrillation was associated with age (P<0.001), repeat surgery (P<0.001); diabetes (P<0.001); hypertension (P=0.002); chronic kidney and pulmonary disease (P=0.026 and 0.005 respectively) as well as mitral valve disease (P<0.001); patients presenting with coronary disease (P=0.005) and acute aortic dissection (P=0.021) less frequently had underlying AF in multivariable analysis (Supplementary Table 2). The origin of CS included acute myocardial infarction (36%) acute aortic dissection (22%) and valvular dysfunction (13%). Other etiologies of CS are shown in Figure 2 and Supplementary Table 3. Acute MI mechanical complications (free wall rupture, papillary muscle rupture, ventricular septal defect and left ventricle aneurysm) constituted 6.9% of cardiogenic shock causes (Figure 2).

**Figure 2.**
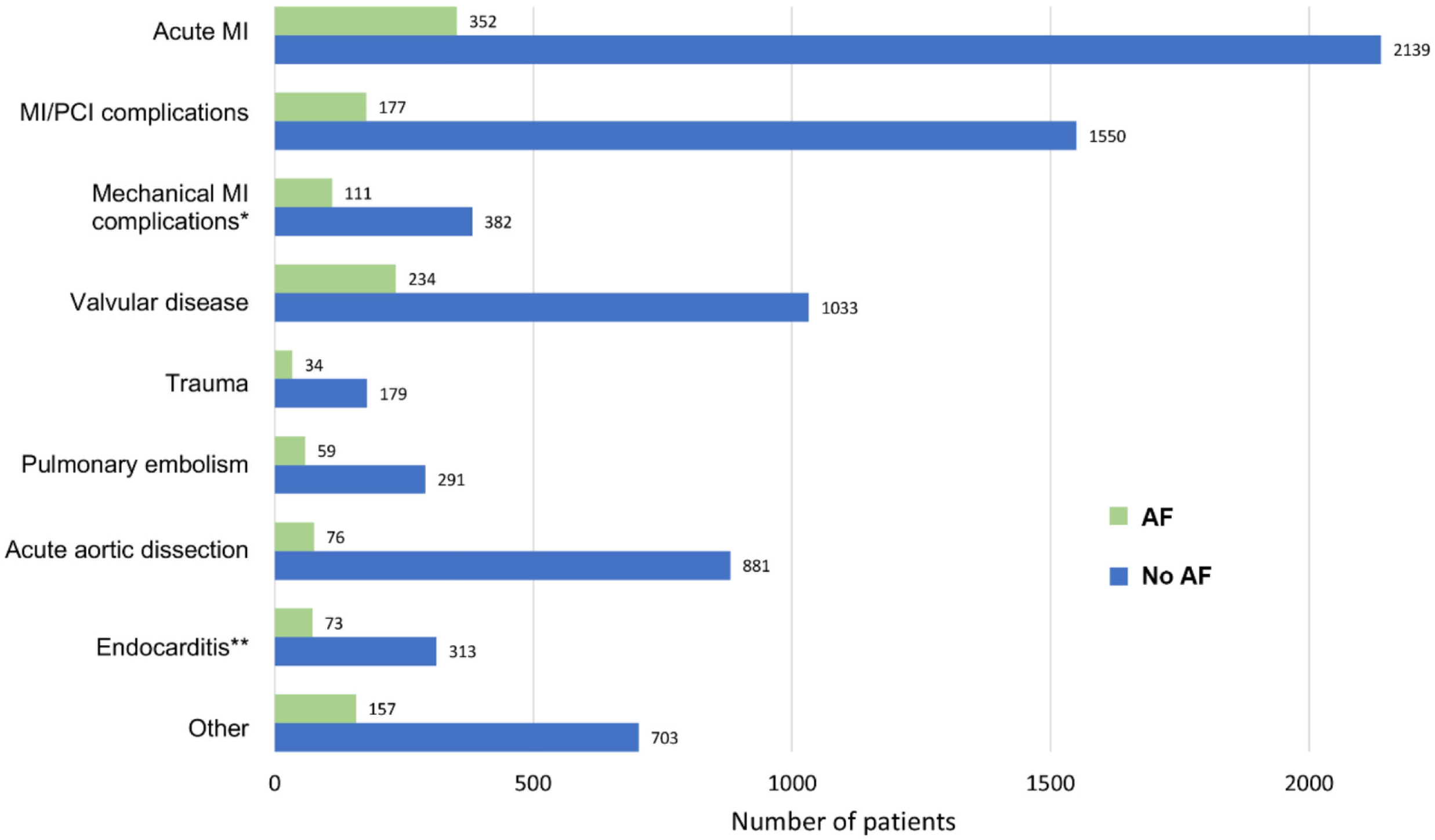
Causes of cardiogenic shock. AF, atrial fibrillation; MI, myocardial infarction.

Mechanical circulatory support was used preoperatively in 920 (21.4%) patients, and most commonly included intra-aortic balloon pump in 876 (18.1%) patients; followed by ECMO in 78 (3.0%) patients and VAD in 97 (2.0%).

Coronary artery bypass grafting was most commonly performed procedure [1,594 pts (32.9%)] followed by aortic dissection repair in 957 (19.7%); aortic, mitral and tricuspid valve repair or replacement surgery was performed in 727 (15.0%), 551 (11.4%) and 140 (2.9%) cases respectively. Fifty-eight (1.2%) patients underwent orthotopic heart transplantation while 101 (2.1%) underwent VAD implantation. Median ICU length of stay was 101.5 hours [Interquartile range (IQR): 47.3-213.6] and HLoS among those who survived to discharge 9.2 days (IQR: 5.7-16.6). Surgical data are reported in Supplementary Table 4.

Thirty-day mortality was 35.6%. In-hospital complications are available as Supplementary Table 5. In multivariable analysis, age (P<0.001); repeat surgery (P=0.012); hypertension (P=0.001); chronic kidney disease (P<0.001); peripheral artery disease (P<0.001); mechanical ventilation (P<0.001) and surgical urgency (P<0.001) were associated with long-term mortality (Supplementary Table 6). In an unadjusted analysis, patients with AF had almost 20% higher mortality risk (HR 1.19, 95% CIs 1.06-1.34; P=0.004) (Supplementary Figure 1).

### PS-matched analysis

We performed a propensity score analysis after the exclusion of orthotopic heart transplantation patients. After the PS-matching 597 pairs were identified (Figure 1). Baseline characteristics of the study cohort are summarized in Table 1. Patients with AF had more previous percutaneous coronary artery intervention (13.7% vs 18.9%; P=0.01), whereas no other significant differences regarding the prevalence of cardiovascular risk factors and comorbidities were seen (Table 1, Supplementary Figure 2 - SMD figure Love plot, Supplementary Figure 3 - PS distribution plot). Principal causes of cardiogenic shock are listed in Table 2. We observed no marked differences between AF and no AF patients in terms of CS origin. Little less than 30% of patients in both groups were operated on shortly after MI (6.5% had mechanical AMI complications). In 15% of patients in both groups acute aortic dissection was the indication for emergent surgery, while pulmonary embolism and infective endocarditis accounted for around 10% in each group.

**Table 1.**
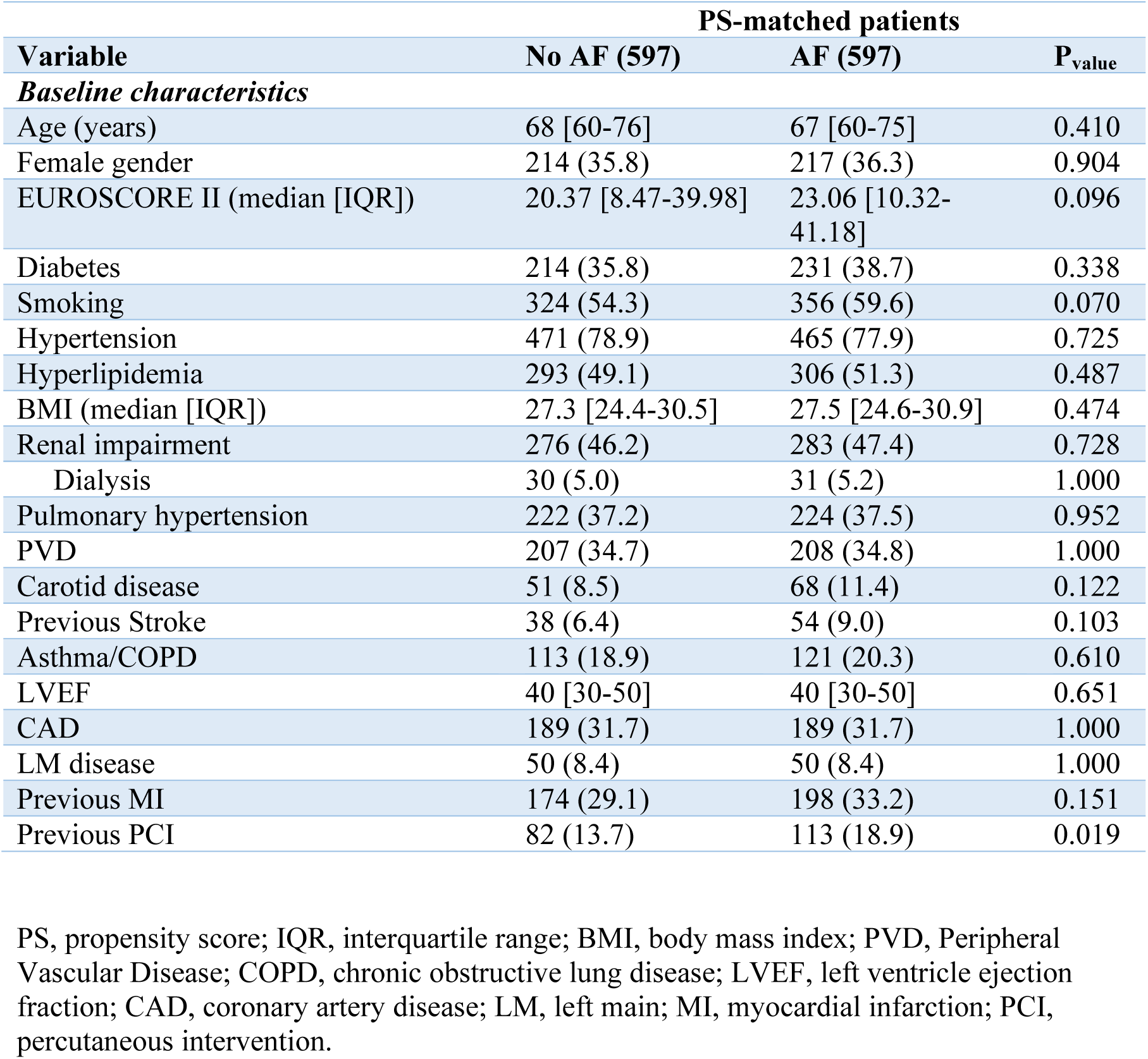
Preoperative characteristics after PS-matching

**Table 2.** Principal causes of cardiogenic shock

|  | PS-matched patients |  |  |
| --- | --- | --- | --- |
|  | No AF (597) | AF (597) | P<sub>value</sub> |
| ***Cause of Cardiogenic Shock*** |  |  |  |
| Acute MI | 178 (29.8) | 174 (29.1) | 0.849 |
| MI Mech Complications | 39 (6.5) | 39 (6.5) | 1.000 |
| VSD | 23 (3.9) | 30 (5.0) | 0.399 |
| PM rupture | 7 (1.2) | 6 (1.0) | 1.000 |
| Free wall rupture | 4 (0.7) | 2 (0.3) | 0.687 |
| Ventricular aneurysm | 12 (2.0) | 6 (1.0) | 0.234 |
| Unstable Angina | 39 (6.5) | 20 (3.4) | 0.016 |
| PE | 56 (9.4) | 69 (11.6) | 0.257 |
| Valvular disease | 144 (24.1) | 144 (24.1) | 1.000 |
| Infective Endocarditis | 69 (11.6) | 70 (11.7) | 1.000 |
| AAD | 88 (14.7) | 78 (13.1) | 0.452 |
| Trauma | 29 (4.9) | 30 (5.0) | 1.000 |
| Other | 109 (18.3) | 140 (23.5) | 0.033 |

PS, propensity score; MI, myocardial infarction, VSD, ventricular septum defect; PM, papillary muscle; PE, pulmonary embolism; AAD, acute aortic dissection

Surgical data are listed in Table 3. There was a trend towards higher prevelance of hemodynamic instability, defined as the use of iv inotropes (65.3% versus 70.4%; P=0.072) in the AF group. Coronary artery bypass grafting (23.5 %) and mitral valve procedure (21.6%) were the most commonly performed procedures without significant differences between AF and no-AF patients. In the AF group, the tricuspid valve procedures (4% versus 7.2%; P=0.023) and surgical pulmonary embolectomy rates (1% versus 3%; P=0.021) were higher. Among patients with AF concomitant cardiac ablation was performed in 6 patients (1%) and left atrial appendage (LAA) closure in 12 (2%).

**Table 3.** Operative characteristics after PS-matching

| Variable | PS-matched patients |  |  |
| --- | --- | --- | --- |
|  | No AF (597) | AF (597) | P<sub>value</sub> |
| ***Procedural characteristics*** |  |  |  |
| iv. inotropes | 390 (65.3) | 420 (70.4) | 0.072 |
| iv. nitrates | 228 (38.2) | 231 (38.7) | 0.905 |
| IABP | 96 (16.1) | 95 (15.9) | 1.000 |
| VAD pre-op | 12 (2.0) | 12 (2.0) | 1.000 |
| ECMO | 10 (1.7) | 19 (3.2) | 0.131 |
| Mechanical ventilation | 223 (37.4) | 225 (37.7) | 0.952 |
| ***Urgency*** |  |  |  |
| Urgent | 158 (26.5) | 185 (31.0) | 0.096 |
| Emergent | 287 (48.1) | 273 (45.7) | 0.451 |
| Salvage | 152 (25.5) | 139 (23.3) | 0.419 |
| ***Surgery*** |  |  |  |
| CPB time* (median [IQR]) | 131 [95-189] | 131 [95-182] | 0.712 |
| X-clamp time* (median [IQR]) | 80 [51-106] | 79 [53-112] | 0.726 |
| Redo surgery | 63 (10.6) | 103 (17.3) | 0.001 |
| MVR | 123 (20.6) | 135 (22.6) | 0.439 |
| AVR | 106 (17.8) | 97 (16.2) | 0.538 |
| TVPR | 24 (4.0) | 43 (7.2) | 0.023 |
| CABG | 150 (25.1) | 134 (22.4) | 0.308 |
| Post inf VSD | 15 (2.5) | 26 (4.4) | 0.111 |
| Aorta repair | 110 (18.4) | 101 (16.9) | 0.544 |
| ECMO (central cannulation) | 30 (5.0) | 36 (5.7) | 0.527 |
| VAD implantation | 7 (1.2) | 21 (3.5) | 0.012 |
| Trauma | 29 (4.9) | 30 (5.0) | 1.000 |
| Tumor | 2 (0.3) | 2 (0.3) | 1.000 |
| Embolectomy | 6 (1.0) | 18 (3.0) | 0.021 |
| Other | 107 (17.9) | 129 (21.6) | 0.127 |

\*missing data
PS, propensity score; IABP, intra-aortic balloon pump; iv., intravenous; OPCAB, Off-Pump Coronary Artery Bypass; CPB, cardiopulmonary bypass; LAAO, left atrial appendage occlusion; VSD, ventricular septal defect; SD, Standard Deviation.

The use of mechanical circulatory support (pre-operative ventricular assist device (2% versus 2%; P=1.000) and extracorporeal membrane oxygenation (1.7% versus 3.2%; P=0.131) was similar in both groups.

In hospital outcomes are reported in Table 4. The major postoperative outcomes: severe bleeding requiring re-thoracotomy, respiratory failure, neurological and gastrointestinal complications; superficial and deep sternal wound infection and the use of ECMO and intra-aortic balloon pump was similar in both groups. In the PS-matched analysis, total 30-day mortality was 33.6% and was numerically higher in AF group (34.7 vs 32.5%; P=0.462) with incidence rates varying across type of surgical procedures; AAD repair had highest (41.8%), followed by AVR/r (38.9%), CABG+valve (38.6%), TVR/r 36.8%, multivalve surgery 36.2% and mitral valve procedures (35.3%), without significant differences between AF and No AF groups but CABG group (42.7 vs 26.9%; P=0.005) in favor of no AF (Figure 3). Median follow-up was 4.6 years (max.10.0 years) and it was 100% complete for the mortality outcome; AF remained associated with worse survival (HR 1.19, 95% CI 1.00-1.40; P=0.045) (Figure 4) at long term.

**Table 4.** In-hospital outcomes after PS-matching

| *Variable* | **PS-matched patients** |  |  |
| --- | --- | --- | --- |
|  | **No AF (597)** | **AF (597)** | **P<sub>value</sub>** |
| 30-day mortality | 194 (32.5) | 207 (34.7) | 0.462 |
| Cardiac tamponade and/or rethoracotomy for bleeding | 71 (11.9) | 82 (13.7) | 0.387 |
| Respiratory failure | 95 (15.9) | 117 (19.6) | 0.112 |
| Neurologic complications | 45 (7.5) | 46 (7.7) | 1.000 |
| Multiorgan failure | 132 (22.1) | 127 (21.3) | 0.779 |
| Gastrointestinal complications | 14 (2.3) | 21 (3.5) | 0.303 |
| Acute kidney failure and/or dialysis | 74 (12.4) | 85 (14.2) | 0.394 |
| Superficial sternal wound infection | 9 (1.5) | 10 (1.7) | 1.000 |
| Deep sternal wound infection | 4 (0.7) | 7 (1.2) | 0.547 |
| PPI | 3 (0.5) | 5 (0.8) | 0.726 |
| ECMO | 48 (8.0) | 61 (10.2) | 0.228 |
| ECMO days | 5 [1, 7] | 5 [3, 13] | 0.261 |
| IABP | 71 (11.9) | 74 (12.4) | 0.859 |

PS, propensity score; AF, atrial fibrillation; PPI, permanent pacemaker implantation; ECMO, extracorporeal membrane oxygenation; IABP, intra-aortic balloon pump.

**Figure 3.**
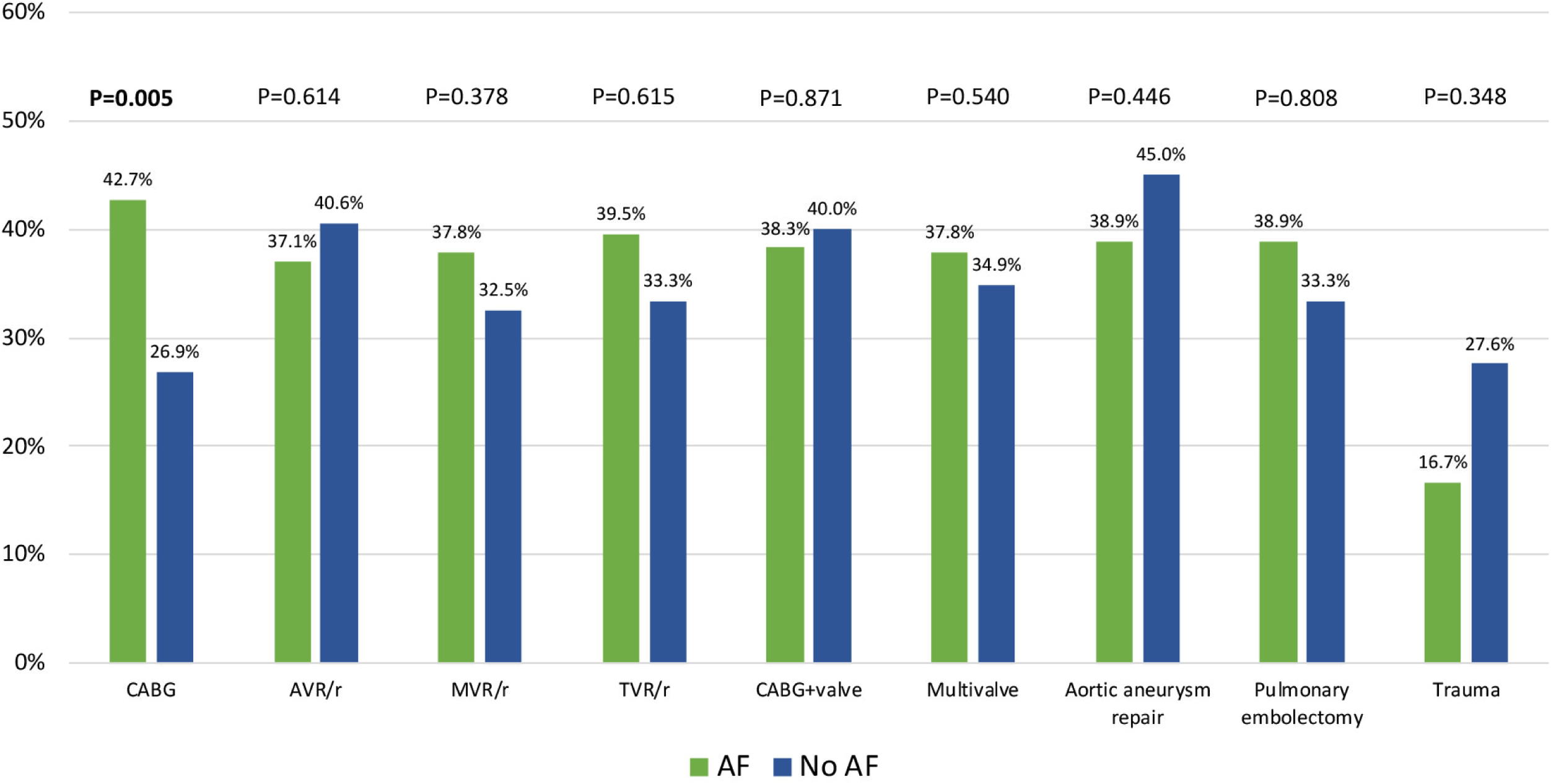
30-day mortality in AF and no AF groups according to the type of surgery. CABG, Coronary Artery Bypass Grafting; AVR, aortic valve replacement; MVR, mitral valve replacement; TVR/r, tricuspid valve replacement/repair.

**Figure 4.**
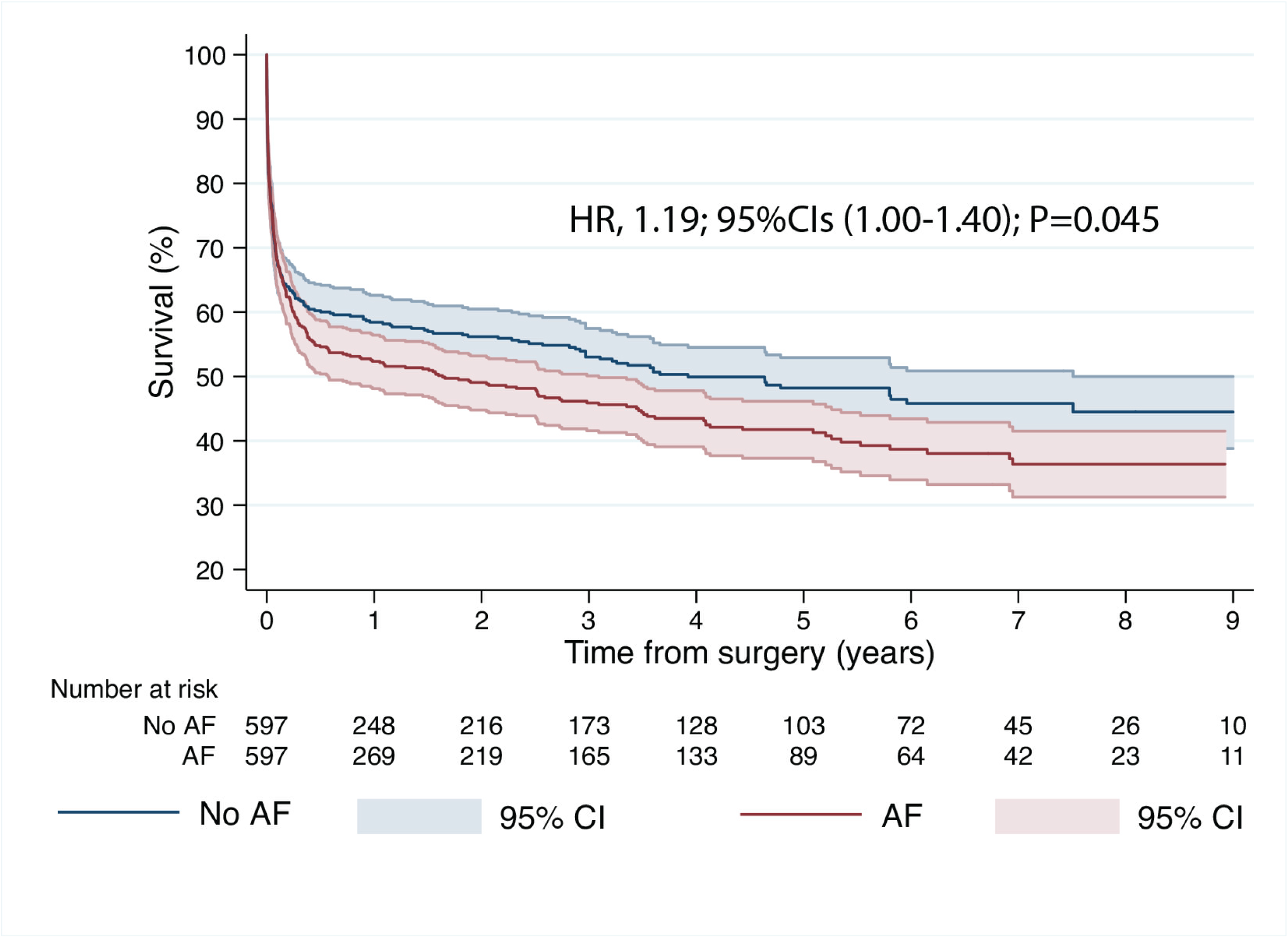
Adjusted Kaplan–Meier curve displaying survival according to presence or absence of AF. AF, atrial fibrillation; HR, hazard ratio; CI, Confidence Intervals.

Proportional hazard assumption was not violated (P=0.439) as also graphically assessed (Supplementary Figure 4 and 5). In the subgroup analysis, the harmful effect of AF on long-term mortality was seen in patients initially presenting with unstable coronary artery disease (P=0.024) and valvular disease (P=0.030), in particular IE (P=0.007). Supplementary Figure 6.

## Discussion

To the best of our knowledge, this is the first study from a large national inpatient database to analyze the prognostic impact of underlying AF in various setting of CS requiring heart surgery. As major findings, history of AF strongly impacts 1) the survival at 30 days driven by reduction of mortality in patients undergoing CABG surgery and 2) and is associated with higher long-term mortality regardless of the etiology of CS. 3) postoperative complications were similar in patients with and without documented AF during index hospital stay; furthermore, 4) concomitant ablation of AF and closure of left atrial appendage are rarely performed during cardiac procedure for CS.

Atrial fibrillation is the most common cardiac arrhythmia in the general population and a lifetime risk of > 20% after the age of 55 [11,26]. Its prevalence is estimated to at least double with ageing population [11]. Stroke is the most feared complication in patients with AF, however it also impacts on clinical outcome in specific clinical conditions such as AMI and heart failure or following cardiac surgery procedure [13–17, 27]. As many as 28% of the patients admitted for heart surgery procedure present with AF with increasing rates depending on the presence of valvular dysfunction and extent of the cardiac disease [28]. AF is a well-known marker of high-risk patients and a predictor of postoperative complications including mortality, postoperative stroke, renal failure, prolonged ventilation, reoperation, and deep sternal wound infection [13]. Patients with preoperative diagnosis of AF also experience a higher adjusted long-term risk of all-cause death and of a cumulative risk of stroke and systemic embolism compared to those without [13].

### AF and CS post AMI

The prognostic impact of AF in the setting of CS complicating AMI has mostly been reported after percutaneous procedures. From the IABP-SHOCK II trial (600 patients enrolled, 169 with AF versus 431 without), there were no significant differences with respect to mortality at 30 days and 12 months between patients with and without AF [29]. Similarly, the rates of recurrent MI, repeat revascularization, and stroke did not differ between groups. The authors did not observe any interaction between the impact of IABP on clinical outcome and the prevalence of AF. Reflecting the above were the findings reported in a sub-analysis of the Culprit Lesion Only PCI versus Multivessel PCI in Cardiogenic Shock trial [15]. The study included 686 patients (142 with AF history on admission, or newly detected AF during index hospitalization) and AF was not a significant predictor of 30-day and 1-year all-cause mortality. However, patients with AF already on admission (90 of 142), showed higher all-cause mortality at 30 days (58% versus 37%; P=0.02) and 1 year (63% versus 39%; P=0.004) compared with patients with newly detected AF during hospital stay. Furthermore, AF was associated with longer time to hemodynamic stabilization (4 versus 3 days; P=0.04) at 30 days. In another PS-matched study from NIS registry including 840 patients (420 with AF) who underwent PCI while on percutaneous VAD (Impella^®^) because of CS complicating AMI, all-cause in-hospital mortality rates between the two groups were similar (40.5% vs 36.7%, p=0.245). However, the AF group experienced a significantly higher rate of postprocedural respiratory complications (9.5% vs 4.8%; P=0,007), fewer routine discharges (13.1% vs 30.2%; P<0.001) and more frequent transfers to other healthcare facilities (27.3% vs 17.8%; P<0.001). The mean LOS (12 vs 9 days; P<0.001) and hospital charges ($308,478 vs $277,982; P=0,008) were higher in the AF group [16].

### AF and CS - surgical strategy

The impact of preoperative AF in patients requiring heart surgery for CS is poorly investigated and reported. In our study, CAD remains the major cause of CS and CABG remains the most common surgical treatment, respectively. The current guidelines do not exclude a role for emergency CABG that is usually regarded as the last resource and only in a very limited percentage of patients [30]. Patients undergoing isolated CABG for CS suffer up to 20% higher mortality rates comparing to those without and this occurs also with milder degrees of CS [31,32]. In one recent analysis from the STS database, of the 5,259 patients with AMI complicated by CS who underwent CABG during the study period, 665 (12.6%) patients had AF which in a multivariable logistic regression analysis was associated with increased operative mortality (HR 1.44, 95% CI [1.18–1.77]; P<0.001) [33].

Recent reports on surgery for mitral papillary muscle rupture and CS from the Japan cardiovascular surgery database (196 patients, 140 CS) and STS database (1,342 patients, 759 CS) do not address this issue (the former) or describe no impact of preoperative arrhythmias as predictors of operative mortality in multivariable logistic regression model (the latter) [34, 35]. Sagakuchi et al. identified 1,397 patients undergoing surgical repair of post-MI VSD (61.5% CS) from the national Japanese database and concluded that preoperative AF was not a significant prognostic factor (HR 0.79, 95% CI [0.50-1.23]; P=0.29 in the multivariable analysis) [supplementary reference 1]. Similarly, no relationship was observed between the prevalence of the AF and survival in the UK National Adult Cardiac Surgery Audit of post-infarct ventricular septal defect repair (5.0% among survivors, 5.9% among non-survivors; P=0.6) [supplementary reference 2]. Correspondingly, in our study, we did not observe differences in survival in the MI mechanical complications subgroup. However, we noted a significant relationship among patients with different CS etiology, particularly CAD and valvular decease.

One interesting finding of the current analysis is the low utilization rate of MCS devices in patients with CS in anticipation of surgical treatment. In the setting of CS, temporary MCS can help to stabilize patients and grant time for decision-making about the definitive management [31]. In a recent STS report, AF occurrence in patients with AMI and CS undergoing CABG was higher in the MCS group suggesting a further negative hemodynamic impact of this arrhythmia [31]. In our analysis only 19.0% patients received MCS, and these most commonly included IABP (18.1%); followed by ECMO in 78 (3.0%) patients and VAD in 97 (2.0%). What is reflected in the present analysis is the approach to rush the patient to the OR and stabilize the condition with CPB in most cases rather than stabilize the patient first in the ICU.

### “Anti-AF” approaches

This study shows that ablation of AF or LAAO during heart surgery for patients in CS is very seldom performed. From the 2020 STS report, only 18 patients among 1342 (1.3%) that underwent mitral valve surgery for ischemic papillary muscle rupture received ablation and in three major randomized trial on LAA closure (LAOS I – III) non-elective surgical cases were excluded by the study design [supplementary references 2-5] . Conditions related to CS requiring surgery are demanding and challenging operations and it is perfectly understandable that management of the cause of CS should be the priority. The current analysis could not address AF surgical management; yet, because of lower mortality in the no-AF matched, it may suggest there is a potential to reduce both early and long-term mortality when AF is addressed as well. Indeed, previous observational studies suggested similarly lower risk of long-term mortality in patients undergoing surgical ablation concomitant to CABG w/wo valvular procedure in patients in critical condition, with pre-op IABP and on pharmacological inotropic support [20].

### Limitations

There are certain limitations to the current retrospective study that need to be acknowledged; firstly, the registry did not collect, at the time of conception, the data regarding long-term outcomes other than all-cause mortality e.g. long-term stroke, rehospitalization for heart failure, repeat revascularization, re-do surgery and other procedures e.g. catheter ablation or PCI; these could further enhance the registry and might have influenced the remote outcome as well. Secondly, certain detailed baseline and operative data such as AF type and duration were not collected by the registry; information on the timing of interventions, delay to surgery, duration of pre-op IABP, doses of inotropes and certain characteristics of mechanical ventilation and other ICU variables are missing. Finally, while PSM accounted for the variables included in the EuroSCORE II and other surgically relevant characteristics minimizing selection bias in an attempt to even baseline patients’ characteristics, unmeasured biases and confounders may remain, in particular in the setting of cardiogenic shock, making the association between AF and higher mortality in cardiogenic shock valid only to the extent an analysis of a non-RCTs study allows. On the other hand, multivariable analyses fully support the concept of AF as a hallmark of worse baseline condition and higher risk independently associated with worse prognosis both at early and long-term follow-up. The optimal timing of surgical intervention in patients with CS that could benefit of preoperative MCS is a matter of further debate not addressed by this study.

## Conclusions

Among patients with CS referred for cardiac surgery, history of AF was a significant risk factor for both early and long-term mortality. Addressing AF by concomitant ablation and/or left atrial appendage closure at the time of surgery may be considered to reduce thromboembolic risk and worsening of heart failure even in these highest risk patients. However, additional and dedicated studies investigating patients in CS and affected by preoperative AF should be undertaken to carefully analyze the actual impact and related therapeutic treatment to abolish such a cardiac arrhythmia in this peculiar hemodynamic setting.

## Clinical Perspectives

### Core clinical competencies

The current work is the first to address underlying atrial fibrillation in cardiogenic shock patients undergoing cardiac surgery. After adjusting for baseline variables, AF remains significantly linked to mortality even in this highest surgical risk group.

### Translational outlook implications

Up to 10% of patients undergoing heart surgery present with underlying AF. Those undergoing elective procedures, have surgical ablation and/or left atrial appendage occluded to a degree depending on the type of surgery and local reimbursement policy. Patients operated in cardiogenic shock, have their AF seldom addressed. Future studies should look at anti-AF and anti-thrombotic strategies in this particular population.

## Data Availability

The datasets generated during and/or analyzed during the current study are available from the corresponding author on reasonable request.

## Acknowledgement

We acknowledge the contribution of late Professor Doctor Marian Zembala (1950-2022), pioneer in cardiac surgery, Head of Silesian Centre for Heart Diseases in Zabrze and the Minister of Health, in the conception and design of the KROK registry.

KROK investigators are listed in the supplement.

Work by Thoracic Research Centre (www.trc.org.pl)

## Author contribution statement

Conceptualization: MK, GMR; data curation: MK, GMR, MP; formal analysis: MK, MP; funding acquisition: NA; investigation: all authors; methodology: all authors; project administration: MK,PS; resources: MK, PS; software: MK; supervision: MK, PS; validation: all authors; visualization: MK, MP; writing-original draft: all authors; writing-review and editing: all authors. Other: registry administration: ZT.

